# A Comparative Study: Diagnostic Performance of ChatGPT 3.5, Google Bard, Microsoft Bing, and Radiologists in Thoracic Radiology Cases

**DOI:** 10.1101/2024.01.18.24301495

**Authors:** Yasin Celal Gunes, Turay Cesur

**Affiliations:** Department of Radiology, Kirikkale Yuksek Ihtisas Hospital, Kirikkale, Türkiye; Department of Radiology, Mamak State Hospital, Ankara, Türkiye

**Keywords:** chatGPT, bard, bing, large language model, thoracic radiology

## Abstract

**Purpose:** To investigate and compare the diagnostic performance of ChatGPT 3.5, Google Bard, Microsoft Bing, and two board-certified radiologists in thoracic radiology cases published by The Society of Thoracic Radiology.

**Materials and Methods:** We collected 124 “Case of the Month” from the Society of Thoracic Radiology website between March 2012 and December 2023. Medical history and imaging findings were input into ChatGPT 3.5, Google Bard, and Microsoft Bing for diagnosis and differential diagnosis. Two board-certified radiologists provided their diagnoses. Cases were categorized anatomically (parenchyma, airways, mediastinum-pleura-chest wall, and vascular) and further classified as specific or non-specific for radiological diagnosis. Diagnostic accuracy and differential diagnosis scores were analyzed using chi-square, Kruskal-Wallis and Mann-Whitney U tests.

**Results:** Among 124 cases, ChatGPT demonstrated the highest diagnostic accuracy (53.2%), outperforming radiologists (52.4% and 41.1%), Bard (33.1%), and Bing (29.8%). Specific cases revealed varying diagnostic accuracies, with Radiologist I achieving (65.6%), surpassing ChatGPT (63.5%), Radiologist II (52.0%), Bard (39.5%), and Bing (35.4%). ChatGPT 3.5 and Bing had higher differential scores in specific cases (P<0.05), whereas Bard did not (P=0.114). All three had a higher diagnostic accuracy in specific cases (P<0.05). No differences were found in the diagnostic accuracy or differential diagnosis scores of the four anatomical location (P>0.05).

**Conclusion:** ChatGPT 3.5 demonstrated higher diagnostic accuracy than Bing, Bard and radiologists in text-based thoracic radiology cases. Large language models hold great promise in this field under proper medical supervision.

---

Natural language processing (NLP) tools, specifically large language models (LLMs), have undergone extensive training on substantial datasets, enabling them to generate highly accurate and human-like texts (1). In contemporary landscapes, AI-driven LLMs, such as ChatGPT 3.5, Google Bard, and Microsoft Bing, have gained significant attention for their potential in addressing complex tasks and improving information accessibility across diverse domains (2). This surge in AI-driven advancements has sparked the proliferation of publications in various medical domains including education, medical writing, and diagnostics (3). Radiology, which plays a pivotal role in this landscape, has contributed significantly to the discourse through studies ranging from refining radiological reports and board-style radiology exams to essential preprocedural patient education in interventional radiology (4–7). The domain of radiology quizzes, focused on patients’ medical histories and imaging findings, is a key area of research, with neuroradiology often under scrutiny for performance comparisons of LLMs (8–10). In the specific realm of thoracic radiology, where diagnoses involve meticulous evaluations of clinical data and imaging findings, applications of artificial intelligence are increasingly observed, notably in lung nodule detection, chest X-ray interpretation, and characterization of pleural plaques (11–13). The ongoing debate over whether artificial intelligence can replace thoracic radiologists remains a topic of discussion (14). In a recent study, the suitability of various LLMs for generating differential diagnoses of common patterns in thoracic radiology was investigated (15). Additionally, another study compared the success of only ChatGPT 3.5 and ChatGPT 4 in solving thoracic radiology cases (16). While the integration of LLMs into the decision-making process in radiology is believed to potentially reduce diagnostic errors, it is noteworthy that there is no study for comparing LLMs’ and radiologists’ diagnostic performance in thoracic radiology cases (4). The Society of Thoracic Radiology (STR) shares interesting and significant cases related to thoracic radiology on its website every month, featuring real-life scenarios with patients’ medical histories and imaging findings, known as “Case of the Month”.

This study aimed to evaluate and compare the diagnostic performance of ChatGPT 3.5, Google Bard, Microsoft Bing, and two board-certified radiologists in thoracic radiology cases published monthly by the STR, with a specific focus on the diagnostic performance across a wide variety of diseases.

## MATERIALS and METHOD

### Study Design

This cross-sectional observational study compared LLMs (ChatGPT 3.5, Google Bard, and Bing) and the responses of board-certified radiologists in solving thoracic radiology cases. The study did not require ethics committee approval as it relied solely on published online cases. Its design conformed to the principles articulated in the Standards for Reporting Diagnostic Accuracy Studies (STARD) statement and the Declaration of Helsinki (17).

### Data Collection

Since 2012, the Thoracic Society of Radiology has published monthly cases in the “Case of the Month” category on its website (https://thoracicrad.org). These cases, which encompass patient medical history, imaging findings, diagnoses, differential diagnoses, and discussion sections, are openly accessible to the public. A total of 145 cases were reviewed between March 2012 and December 2023. Ten cases were excluded because of insufficient explanations of imaging findings in the “Findings” section; five cases were omitted where the diagnosis was already provided in the same section; four cases were inaccessible due to technical issues on the website; and two cases were disregarded due to inadequate clinical information in the “History” section. This resulted in the inclusion of 124 cases in our analysis.

The medical history of each patient was extracted from the “History” section located above the initial image. Imaging findings were obtained from the explanations in the “Findings” section, and additional images were evaluated by radiologists in the “Additional Images” section. To ensure unbiased evaluation, radiologists and LLMs did not use information from the “CXRHint” section. An overview of the workflow is shown in Figure 1.

**Figure 1.**
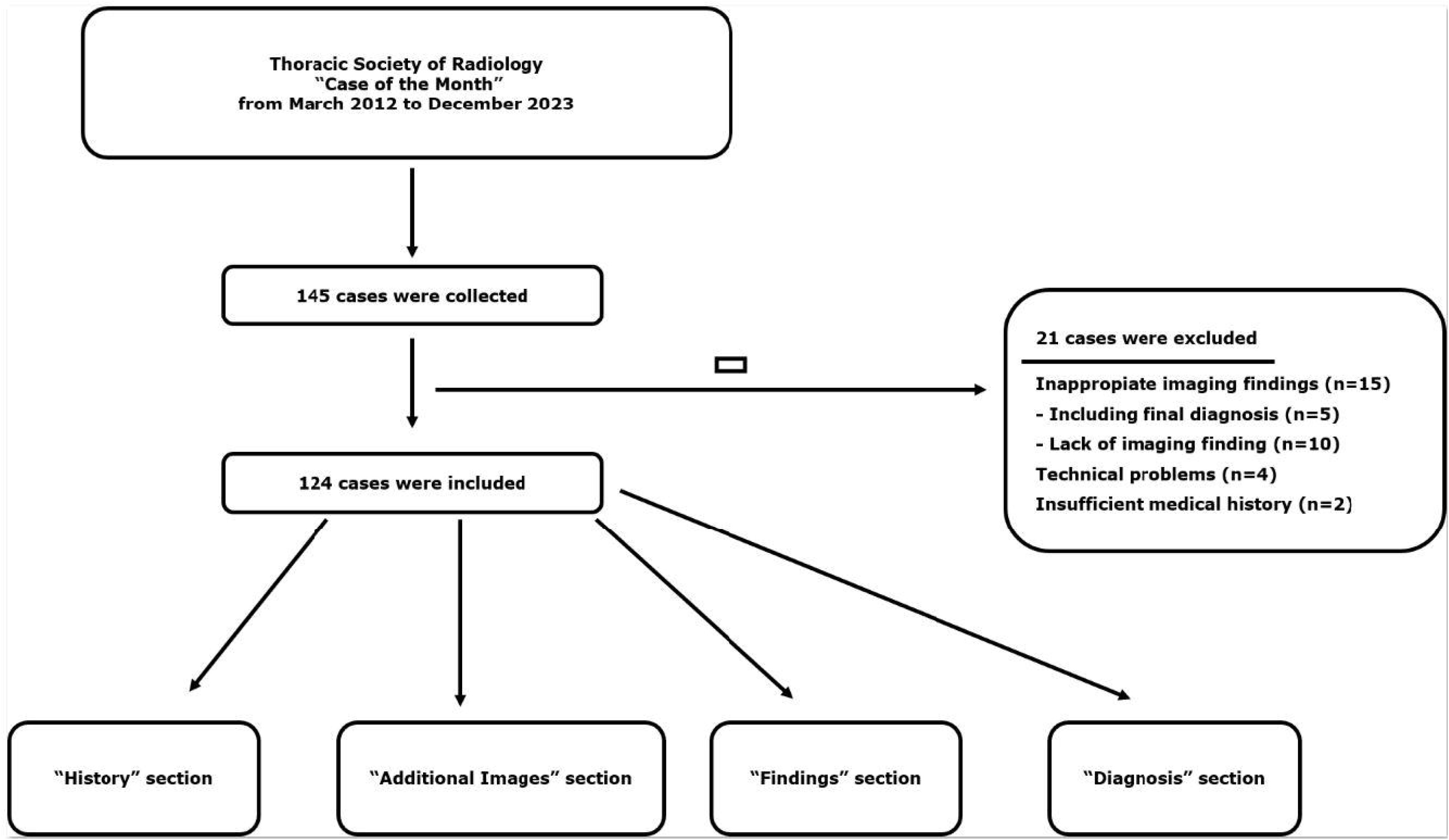
The workflow of the study.

### Category Classification

Cases were categorized into four broad groups based on the thoracic region: parenchymal, airway, mediastinal-pleural-chest wall, and vascular. Furthermore, cases were subcategorized based on the adequacy of radiological images for patient diagnosis. Cases in which radiological images provided sufficient information for diagnosis were termed specific. In contrast, cases in which radiological images were deemed inadequate and the final diagnosis requiring histopathological confirmation were termed non-specific (see Table, Supplemental Digital Content 1, which demonstrates detailed case-specific information). The diagnostic accuracy and performance of ChatGPT 3.5, Bard, and Bing, as well as the diagnoses of the two radiologists, were compared for each group.

### Design of Input-Output Procedures for LLMs

We initiated the input prompt in our study design as “I am working on a radiology quiz and will provide you with medical history and imaging findings. Act like a professor of radiology, please indicate the most likely answer and mention five differential diagnoses, ranked by likelihood.” This prompt was presented in January 2024 on three distinct platforms with default hyperparameters, OpenAI’s ChatGPT 3.5 (https://chat.openai.com) free research version, Google Bard (https://bard.google.com) Experiment, and Microsoft Bing (https://www.bing.com/) Chat (Balanced), using the GPT-4 architecture.

Subsequently, the patients’ medical history and imaging findings were sequentially added to the same chat session. Each LLM was presented in 124 cases, and responses were meticulously recorded. It is crucial to note that the employed LLMs were not pretrained with a specific command or question set for this study. Each question was posed in a single chat session, without opening a new chat tab for individual inquiries (see Text, Supplemental Digital Content 2, which demonstrates examples of prompts given to LLMs and their responses).

### Radiologists’ Interpretation and LLMs’ Performance Evaluation

Two board-certified radiologists, Y. C. G (Radiologist I)- with 6 years of experience- and T. C (Radiologist II)- with 5 years of experience, independently assessed cases blindly using their respective computers. Radiologists classified their responses as either correct (1) or incorrect (0), with correctness based on predefined diagnostic criteria and accuracy in identifying the findings.

Unlike LLMs, radiologists do not create a list of the differential diagnosis for these questions. In a separate session, the differential diagnosis lists generated by ChatGPT 3.5, Bard, and Bing were evaluated by both radiologists for compatibility with the differential diagnosis lists provided by the Thoracic Society of Radiology website for each case. Consensus was reached through joint assessment.

The scoring (DDxScore) was performed as follows:

● Five points (excellent) when both the diagnosis and differential diagnosis were correct.
● Four points (good) when the diagnosis was correct but the differential diagnosis was incomplete.
● Three points (moderate) when the diagnosis was incorrect but the correct diagnosis was in the differential list.
● Two points (poor) when the diagnosis was incorrect and the differential diagnosis list was incomplete.
● One point (very poor) when both the diagnosis and differential diagnosis were incorrect.

This scoring system allows for a nuanced evaluation of both the accuracy of diagnoses and completeness of the differential diagnosis lists.

### Statistical Analysis

The means, standard deviations, medians, minimums,maximums, frequencies, and percentages were used for the descriptive statistics. The distribution of variables was assessed using the Kolmogorov-Smirnov test. Kruskal-Wallis and Mann-Whitney U tests were used to compare quantitative data. The chi-square test was used to compare qualitative data. SPSS 28.0 was used for statistical analyses, and statistical significance was set at P < 0.05.

## RESULTS

A total of 124 ’Case of the Month’ were included in the study. Among these, 77.4% (n=96) were specific, and 22.6% (n=28) were non-specific in terms of radiological diagnosis. ChatGPT correctly answered 53.2% (n=66) of the 124 cases, Radiologist I answered 52.4% (n=65), Radiologist II answered 41.1% (n=51), Bard correctly answered 33.1% (n=41), and Bing correctly answered 29.8% (n=37) (Figure 1).

For specific questions, Radiologist I correctly answered 65.6% (n=63) of the 96 questions, ChatGPT answered 63.5% (n=61), Radiologist II answered 52.0% (n=50), Bard answered 39.5% (n=38), and Bing answered 35.4% (n=34) (Table 1) (Figure 2). For the 17 questions, at least one LLMs provided the correct answer, whereas both radiologists answered the questions incorrectly. Additionally, there were 36 questions, none of which provided the correct answers. In 11 questions known by at least one radiologist, LLMs failed to answer.

**Figure 2.**
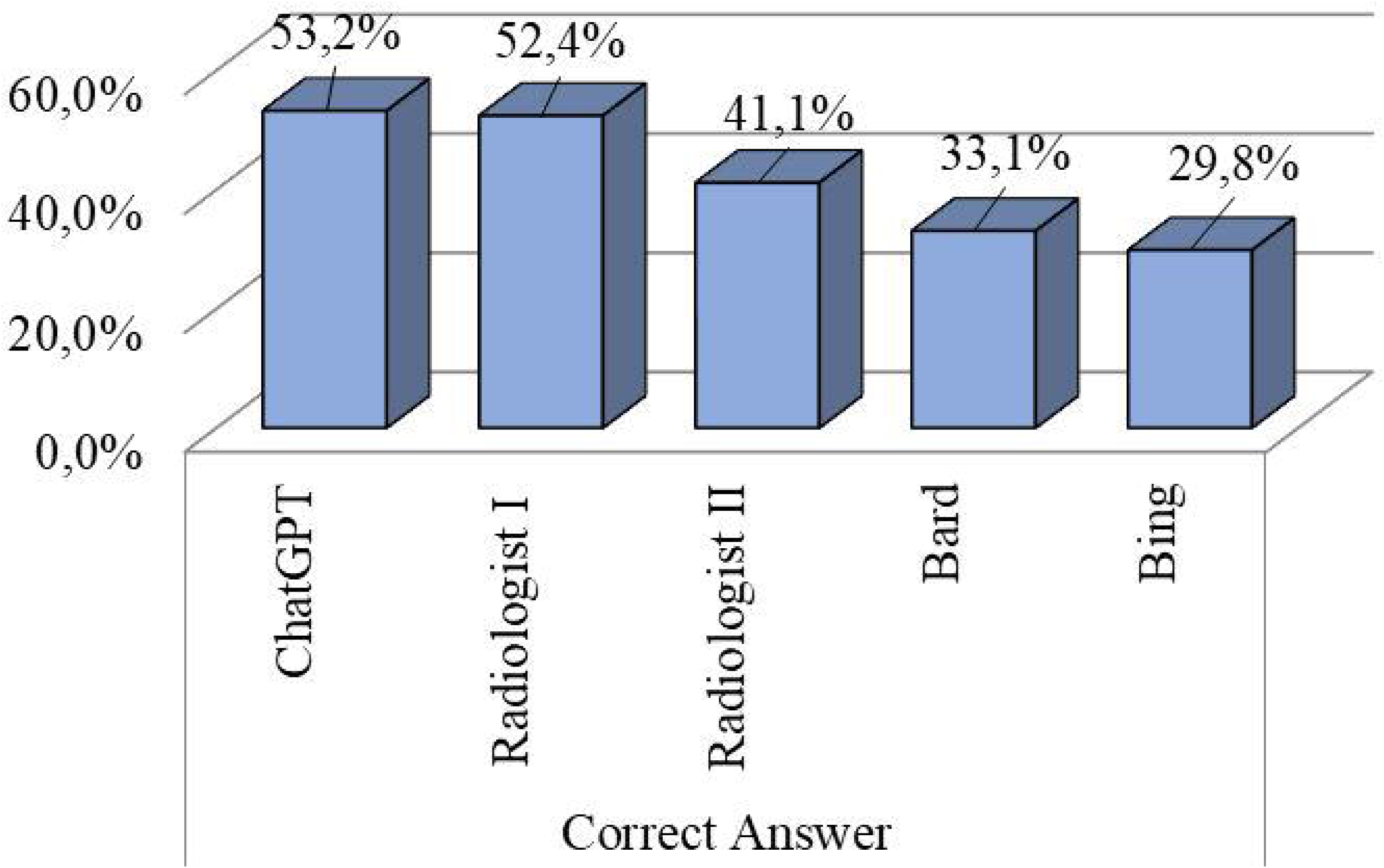
Diagnostic accuracy of large language models and radiologists.

**Figure 3.**
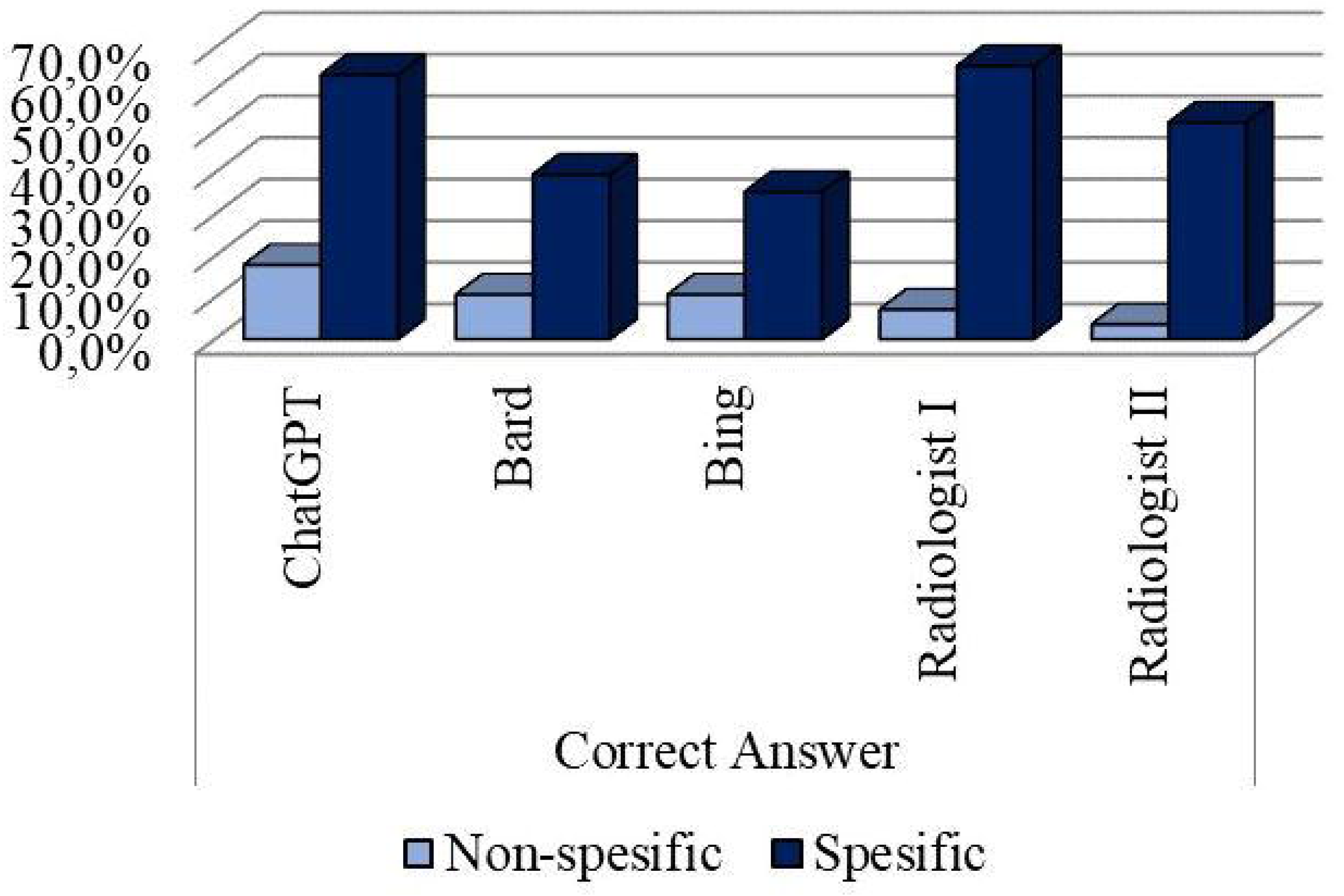
Distribution of correct answers regarding non-specific and specific cases

**Table 1.**
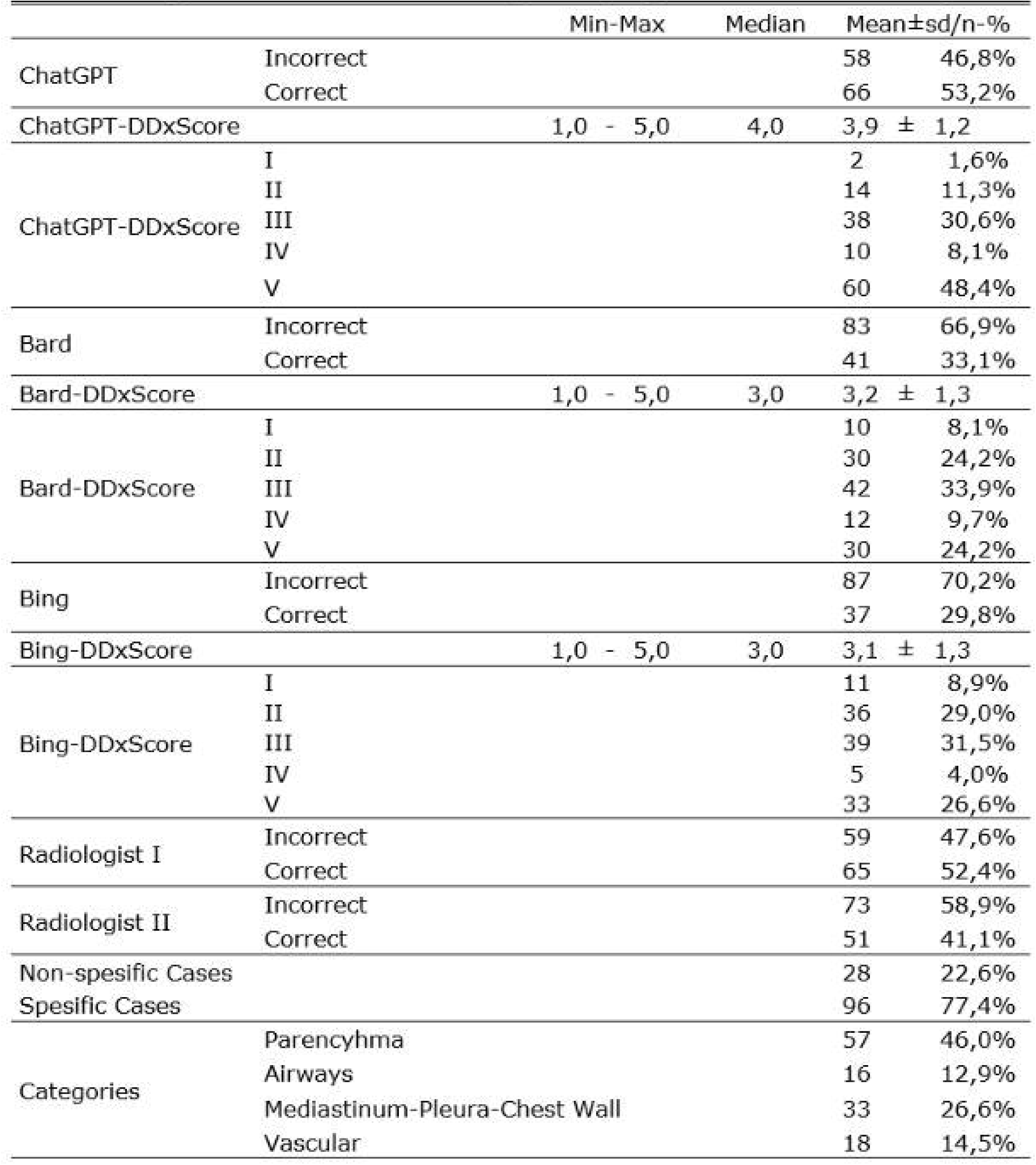
Diagnostic Performance of Large Language Models’ and Radiologists.

|  |  | Min-Max | Median | Mean±sd/n-% |  |
| --- | --- | --- | --- | --- | --- |
| ChatGPT | Incorrect |  |  | 58 | 46,8% |
|  | Correct |  |  | 66 | 53,2% |
| ChatGPT-DDxScore |  | 1,0 - 5,0 | 4,0 | 3,9 ± 1,2 |  |
| ChatGPT-DDxScore | I |  |  | 2 | 1,6% |
|  | II |  |  | 14 | 11,3% |
|  | III |  |  | 38 | 30,6% |
|  | IV |  |  | 10 | 8,1% |
|  | V |  |  | 60 | 48,4% |
| Bard | Incorrect |  |  | 83 | 66,9% |
|  | Correct |  |  | 41 | 33,1% |
| Bard-DDxScore |  | 1,0 - 5,0 | 3,0 | 3,2 ± 1,3 |  |
| Bard-DDxScore | I |  |  | 10 | 8,1% |
|  | II |  |  | 30 | 24,2% |
|  | III |  |  | 42 | 33,9% |
|  | IV |  |  | 12 | 9,7% |
|  | V |  |  | 30 | 24,2% |
| Bing | Incorrect |  |  | 87 | 70,2% |
|  | Correct |  |  | 37 | 29,8% |
| Bing-DDxScore |  | 1,0 - 5,0 | 3,0 | 3,1 ± 1,3 |  |
| Bing-DDxScore | I |  |  | 11 | 8,9% |
|  | II |  |  | 36 | 29,0% |
|  | III |  |  | 39 | 31,5% |
|  | IV |  |  | 5 | 4,0% |
|  | V |  |  | 33 | 26,6% |
| Radiologist I | Incorrect |  |  | 59 | 47,6% |
|  | Correct |  |  | 65 | 52,4% |
| Radiologist II | Incorrect |  |  | 73 | 58,9% |
|  | Correct |  |  | 51 | 41,1% |
| Non-spesific Cases |  |  |  | 28 | 22,6% |
| Spesific Cases |  |  |  | 96 | 77,4% |
| Categories | Parencyhma |  |  | 57 | 46,0% |
|  | Airways |  |  | 16 | 12,9% |
|  | Mediastinum-Pleura-Chest Wall |  |  | 33 | 26,6% |
|  | Vascular |  |  | 18 | 14,5% |

In the specific subgroup, the diagnostic accuracy of ChatGPT was significantly higher than that in the non-specific subgroup (P<0.001). Similarly, the ChatGPT-DDxScore demonstrated significantly higher performance in the specific subgroup than in the non-specific subgroup (P<0.001).

For Bard, diagnostic accuracy was markedly higher in the specific subgroup than in the non-specific subgroup (P=0.004). However, there was no significant difference in the Bard-DDxScore between the specific and non-specific subgroups (P=0.144).

For Bing, the diagnostic accuracy in the specific subgroup was significantly higher than that in the non-specific subgroup (P=0.012). Additionally, in the specific subgroup, the Bing-DDxScore was significantly higher than that in the non-specific subgroup (P=0.004).

Radiologist I showed significantly higher diagnostic accuracy in the specific subgroup than in the non-specific subgroup (P<0.001). Similarly, Radiologist II demonstrated significantly higher diagnostic accuracy in the specific subgroup than in the non-specific subgroup (P<0.001) (Table 2).

**Table 2.**
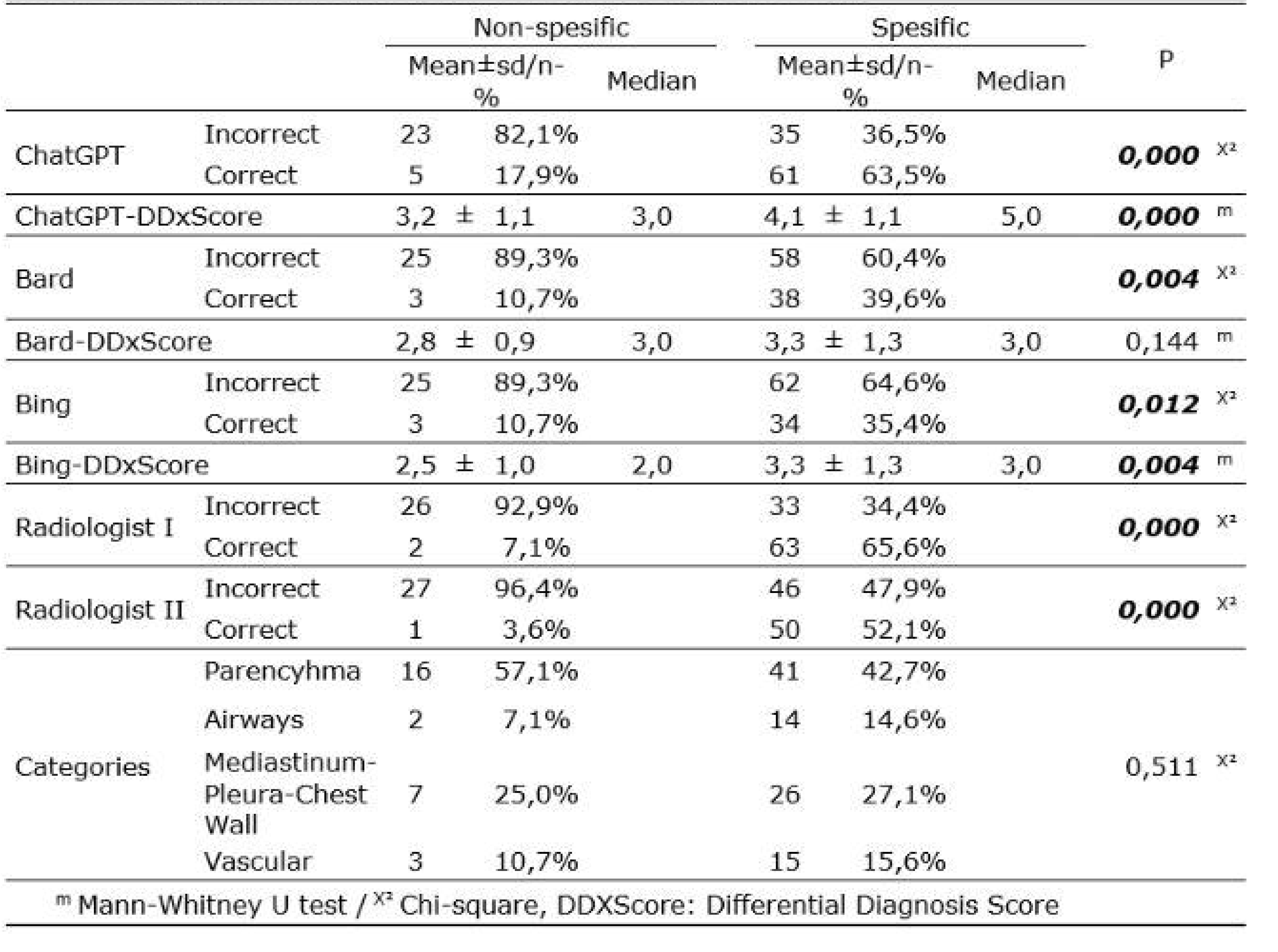
Diagnostic accuracy and classification by cases type.

The parenchyma, airways, mediastinum, and vascular groups did not demonstrate a significant difference in diagnostic accuracy between the ChatGPT, Bard, Bing, Radiologist I, and Radiologist II groups (P>0.05). Similarly, within the parenchyma, airways, mediastinum, and vascular groups, no significant differences were observed in the ChatGPT-DDxScore, Bard-DDxScore, or Bing-DDxScore (P>0.05) (Table 3).

**Table 3.** Diagnostic accuracy and classification by anatomical location.

|  |  |  | Parenchyma |  | Airways |  | Mediastinum-Pleura-Chest Wall |  | Vascular |  | P |
| --- | --- | --- | --- | --- | --- | --- | --- | --- | --- | --- | --- |
| ChatGPT | Incorrect | n-% | 29 | 50,9% | 8 | 50,0% | 14 | 42,4% | 7 | 38,9% | 0,764 <sup>x2</sup> |
|  | Correct | n-% | 28 | 49,1% | 8 | 50,0% | 19 | 57,6% | 11 | 61,1% |  |
| ChatGPT-DDxScore |  | Mean±sd | 3,7 ± 1,3 |  | 4,1 ± 1,1 |  | 4,0 ± 1,2 |  | 4,3 ± 1,0 |  | 0,323 <sup>K</sup> |
|  |  | Median | 3,0 |  | 4,5 |  | 5,0 |  | 5,0 |  |  |
| Bard | Incorrect | n-% | 40 | 70,2% | 11 | 68,8% | 22 | 66,7% | 10 | 55,6% | 0,718 <sup>x2</sup> |
|  | Correct | n-% | 17 | 29,8% | 5 | 31,3% | 11 | 33,3% | 8 | 44,4% |  |
| Bard-DDxScore |  | Mean±sd | 3,1 ± 1,2 |  | 3,1 ± 1,3 |  | 3,2 ± 1,3 |  | 3,5 ± 1,4 |  | 0,656 <sup>K</sup> |
|  |  | Median | 3,0 |  | 3,0 |  | 3,0 |  | 3,5 |  |  |
| Bing | Incorrect | n-% | 41 | 71,9% | 11 | 68,8% | 22 | 66,7% | 13 | 72,2% | 0,954 <sup>x2</sup> |
|  | Correct | n-% | 16 | 28,1% | 5 | 31,3% | 11 | 33,3% | 5 | 27,8% |  |
| Bing-DDxScore |  | Mean±sd | 3,0 ± 1,3 |  | 3,3 ± 1,4 |  | 3,2 ± 1,3 |  | 3,0 ± 1,4 |  | 0,845 <sup>K</sup> |
|  |  | Median | 3,0 |  | 3,0 |  | 3,0 |  | 3,0 |  |  |
| Radiologist I | Incorrect | n-% | 30 | 52,6% | 9 | 56,3% | 13 | 39,4% | 7 | 38,9% | 0,476 <sup>x2</sup> |
|  | Correct | n-% | 27 | 47,4% | 7 | 43,8% | 20 | 60,6% | 11 | 61,1% |  |
| Radiologist II | Incorrect | n-% | 34 | 59,6% | 10 | 62,5% | 18 | 54,5% | 11 | 61,1% | 0,942 <sup>x2</sup> |
|  | Correct | n-% | 23 | 40,4% | 6 | 37,5% | 15 | 45,5% | 7 | 38,9% |  |
| **Questions** |  |  |  |  |  |  |  |  |  |  |  |
| Non-spesific |  | n-% | 16 | 28,1% | 2 | 12,5% | 7 | 21,2% | 3 | 16,7% | 0,511 <sup>x2</sup> |
| Spesific |  | n-% | 41 | 71,9% | 14 | 87,5% | 26 | 78,8% | 15 | 83,3% |  |

<sup>K</sup> Kruskal-wallis (Mann-Whitney U test) / <sup>X<sup>2</sup></sup> Chi-square, DDXScore: Differential Diagnosis Score

## DISCUSSION

In our study, ChatGPT 3.5 demonstrated superior performance (66/124 cases) in answering questions related to thoracic radiology, outperforming radiologists, and other LLMs. The performance of ChatGPT and Radiologist I (65/124 cases) was closely comparable, with ChatGPT 3.5 showing better results in non-specific radiological diagnosis situations (4/25 cases) and Radiologist I performing better in radiologically specific cases (63/99 cases). ChatGPT 3.5 outperformed Radiologist II (51/124 cases), and both radiologists performance surpassed that of Bard (41/124 cases) and Bing (37/124 cases).

Among LLMs, Bing exhibited the lowest diagnostic performance. The specificity of the questions for radiological diagnosis created significant differences in the scoring for both the diagnostic and differential diagnostic assessments of ChatGPT 3.5 and Bing. In contrast, only Bard showed a significant difference in the diagnostic assessment. ChatGPT 3.5 achieved the highest score in differential diagnosis, underscoring its potential clinical impact.

This study revealed that the specificity of radiological diagnosis significantly influences the statistical performance of radiologists. The performance gap between Radiologist I and II was attributed to differences in experience, particularly in rare diseases such as SAPHO and Bird-Hog-Dube syndrome. A factor negatively impacting radiologists’ performance was the static nature of the images, preventing real-time scrolling up and down, and the inability to perform reconstructions, such as MPR.

No significant differences were observed in the performances of radiologists and LLMs based on the category of cases. However, this study identified 17 cases in which radiologists failed to provide accurate diagnoses, thereby highlighting the potential of LLMs for future diagnostic use. LLMs correctly identified cases involving primary pulmonary lymphoma and thymic carcinoid tumors when radiologists were unable to do so. Rare pathologies, such as pulmonary veno-occlusive disease, lymphomatoid granulomatosis, and hepatoid adenocarcinoma of the lung, were observed in both groups, emphasizing the diagnostic challenges posed by these conditions. In 11 questions known to at least one radiologist, LLMs failed to answer, including air embolism, respiratory papillomatosis, and fibrosing mediastinitis.

The divergent performances of Bing, Bard, and ChatGPT 3.5 in our study can be attributed to variations in their designs. Despite Bing and Bard’s Internet access providing a crucial advantage, occasional incorrect responses were identified, possibly linked to information sourced from online platforms rather than scientific literature (18,19). ChatGPT 3.5 demonstrated a distinct advantage owing to its training on a more extensive dataset. However, a notable drawback was its training until September 2021, which limited access to scientific developments thereafter (20).

Sarangi et al. used four distinct LLMs, namely Perplexity, ChatGPT 3.5, Bard, and Bing, to identify ten distinctive patterns observed in thoracic radiology. The subsequent task for these LLMs involved the provision of five differential diagnoses. According to their findings, Perplexity demonstrated the most favorable performance in delivering accurate differential diagnoses, whereas Bard exhibited the least successful performance (15). In contrast to Sarangi et al.’s study, our research demonstrated that ChatGPT 3.5 exhibited the best performance. Notably, our study departs from the introduction of a general pattern for LLMs. Instead, we presented clinical information specific to each case, coupled with the corresponding imaging findings, to mimic real-life scenarios. The task was to make diagnoses and differential diagnoses based on this information. The observed disparity in the performance of the LLMs between the two studies is believed to arise from this fundamental design distinction.

Li et al. demonstrated diagnostic accuracy rates of 40% (14/35 cases) for ChatGPT 3.5 and 46% (16/35 cases) for ChatGPT 4 in thoracic imaging cases, using ’Radiology Diagnosis Please Cases’ to provide patient medical history and imaging findings (16). In contrast, our study with ChatGPT 3.5 revealed a higher diagnostic accuracy of 53.2% (66/124 cases). We attribute these differences to variations in prompts and the inclusion of a broader range of different thoracic radiology cases in our research, covering four distinct anatomical locations. Caruccio et al. demonstrated that varying prompts, ranging from simple to complex, may alter the performance of LLMs (21). We employed freely available and open-access LLMs in our study, leading to the exclusion of ChatGPT-4. However, we believe that our study offers deeper insights by comparing the performance of radiologists with other Language Model Models (LLMs) in both specific and non-specific cases in terms of radiological diagnosis. This comprehensive comparison provided more valuable insights into the radiological knowledge of LLMs.

Rahsepar et al. examined responses to questions related to lung cancer from ChatGPT, Bard, and Bing, identified ChatGPT 3.5 as delivering the most accurate response, achieving a rate of 70.8% (22). Sarangi et al.’s examination of questions resembling FRCR2A, where radiology residents were compared with Bard, Bing, and ChatGPT, showed Bing’s peak performance at 55.33%, whereas Bard lagged behind at 44.17%. Importantly, the performance of radiology residents in their research surpassed that of both LLMs, with Resident I achieving 63.3% and Resident II 57.5% (23).

In Horiuchi et al.’s study, which focused on ChatGPT-4’s responses to diverse questions derived from the ’Case of the Week’ published by the American Journal of Neuroradiology, the diagnostic performance was reported at 50/100 cases. Notably, this study did not establish a clear relationship between anatomical location and accuracy. However, it did observe that ChatGPT-4 has been successfully used to address questions related to CNS and non-CNS tumors (9).

To the best of our knowledge, this study represents an inaugural exploration in the field of thoracic radiology, where clinical information and imaging findings of cases were provided to LLMs, and the results were systematically compared with those of radiologists. This study elucidated the strengths and weaknesses of LLMs in this domain. An additional innovative aspect of this study was the comprehensive evaluation of the performance of LLMs and radiologists across diverse case groups, encompassing parenchymal, airway, mediastinal, pleural, chest wall, and vascular pathologies.

Although this study contributes significantly to the understanding of LLM capabilities in thoracic radiology, it had several limitations. First, the study opted for a methodology in which LLMs were provided with medical information and radiological findings by radiologists for text-based diagnostic assessments, bypassing the direct evaluation of their image recognition capabilities. A recommendation for future studies involves incorporating LLMs with explicit image recognition features, such as ChatGPT-4V, to evaluate their diagnostic performance in interpreting medical images comprehensively. Second, the complexity of cases presented in the “Case of the Month” might not always mirror routine cases encountered in daily thoracic radiology practice. This discrepancy may limit the generalizability of our findings to routine scenarios, emphasizing the need for studies with a broader range of cases. Third, the absence of radiologists with extensive experience in thoracic radiology may have influenced the overall performance comparison between the radiologists and LLMs. Future research should include a diverse group of radiologists, including residents, thoracic radiology fellows, and attending radiologists, for a more comprehensive analysis. Finally, to acknowledge the variability in the diagnostic performance of LLMs using different prompts, a single prompt was used in this study. Future studies should explore the impact of various prompts on the diagnostic performance of LLMs in radiology in order to gain a more nuanced understanding of their capabilities.

In conclusion, this study revealed the superior performance of the ChatGPT 3.5 in thoracic radiology, surpassing that of radiologists and other LLMs. Noteworthy differences in design and training among Bing, Bard, and ChatGPT 3.5 account for their varying outcomes, which holds great promise in this field under proper medical supervision. Future investigations should delve into the image recognition capabilities of LLMs and incorporate diverse case scenarios to provide robust insights into their diagnostic efficacy.

## Supporting information

1-Supplemental Digital Content 1.xls

2-Supplemental Digital Content 2.doc

## Data Availability

Yes,I declared that all data are available upon request.

## ACKNOWLEDGMENTS

The authors would like to thank Ertan Koç for the statistical analysis of this manuscript. The authors used ChatGPT, a language model based on the GPT-3.5 architecture (January 16 Version; OpenAI; https://chat.openai.com/) to revise the grammar and English translation. The content of the publication is entirely the authors’ responsibility, and the authors examined and edited it as necessary.

## List of Supplemental Digital Content

- Supplemental Digital Content 1.xlsx
- Supplemental Digital Content 2.docx

## Abbreviations

DDx: Differential Diagnosis
LLMs: Large language models
NLP: Natural language processing
STARD: Standards for Reporting Diagnostic Accuracy Studies
STR: Society of Thoracic Radiology

## REFERENCES

1. Thirunavukarasu AJ, Ting DSJ, Elangovan K, et al. Large language models in medicine. Nat Med. 2023;29:1930–1940.

2. Seth I, Lim B, Xie Y, et al. Comparing the Efficacy of Large Language Models ChatGPT, BARD, and Bing AI in Providing Information on Rhinoplasty: An Observational Study. Aesthet Surg J Open Forum. 2023;5:ojad084.

3. Sallam M. ChatGPT Utility in Healthcare Education, Research, and Practice: Systematic Review on the Promising Perspectives and Valid Concerns. Healthcare (Basel). 2023;11:887.

4. Bera K, O’Connor G, Jiang S, et al. Analysis of ChatGPT publications in radiology: Literature so far. Curr Probl Diagn Radiol. Published online October 20, 2023.

5. McCarthy CJ, Berkowitz S, Ramalingam V, et al. Evaluation of an Artificial Intelligence Chatbot for Delivery of IR Patient Education Material: A Comparison with Societal Website Content. J Vasc Interv Radiol. 2023;34:1760–1768.

6. Bhayana R, Krishna S, Bleakney RR. Performance of ChatGPT on a Radiology Board-style Examination: Insights into Current Strengths and Limitations. Radiology. 2023;307:e230582.

7. Amin KS, Davis MA, Doshi R, et al. Accuracy of ChatGPT, Google Bard, and Microsoft Bing for Simplifying Radiology Reports. Radiology. 2023;309:e232561.

8. Suthar PP, Kounsal A, Chhetri L, et al. Artificial Intelligence (AI) in Radiology: A Deep Dive Into ChatGPT 4.0’s Accuracy with the American Journal of Neuroradiology’s (AJNR) “Case of the Month”. Cureus. 2023;15:e43958.

9. Horiuchi D, Tatekawa H, Shimono T, et al. Accuracy of ChatGPT generated diagnosis from patient’s medical history and imaging findings in neuroradiology cases. Neuroradiology. 2024;66:73–79.

10. Horiuchi D, Tatekawa H, Oura T, et al. Comparison of the diagnostic performance from patient’s medical history and imaging findings between GPT-4 based ChatGPT and radiologists in challenging neuroradiology cases. medRxiv. 2023.

11. Tejani A, Dowling T, Sanampudi S, et al. Deep Learning for Detection of Pneumothorax and Pleural Effusion on Chest Radiographs: Validation Against Computed Tomography, Impact on Resident Reading Time, and Interreader Concordance. J Thorac Imaging. Published online September 29, 2023.

12. Groot Lipman KBW, Boellaard TN, de Gooijer CJ, et al. Artificial Intelligence-based Quantification of Pleural Plaque Volume and Association With Lung Function in Asbestos-exposed Patients. J Thorac Imaging. Published online November 1, 2023.

13. Abadia AF, Yacoub B, Stringer N, et al. Diagnostic Accuracy and Performance of Artificial Intelligence in Detecting Lung Nodules in Patients With Complex Lung Disease: A Noninferiority Study. J Thorac Imaging. 2022;37:154–161.

14. Eltorai AEM, Bratt AK, Guo HH. Thoracic Radiologists’ Versus Computer Scientists’ Perspectives on the Future of Artificial Intelligence in Radiology. J Thorac Imaging. 2020;35(4):255–259.

15. Sarangi PK, Irodi A, Panda S, et al. Radiological Differential Diagnoses Based on Cardiovascular and Thoracic Imaging Patterns: Perspectives of Four Large Language Models. Indian J Radiol Imaging. Published online December 28, 2023.

16. Li D, Gupta K, Bhaduri M, Sathiadoss P, et al. Comparing GPT-3.5 and GPT-4 Accuracy and Drift in Radiology Diagnosis Please Cases. Radiology. 2024;310:e232411.

17. Bossuyt PM, Reitsma JB, Bruns DE, et al. STARD 2015: An Updated List of Essential Items for Reporting Diagnostic Accuracy Studies. Radiology. 2015;277:826–832.

18. Lim ZW, Pushpanathan K, Yew SME, et al. Benchmarking large language models’ performances for myopia care: a comparative analysis of ChatGPT-3.5, ChatGPT-4.0, and Google Bard. EBioMedicine. 2023;95:104770.

19. Xie Y, Seth I, Hunter-Smith DJ, et al. Investigating the impact of innovative AI chatbot on post-pandemic medical education and clinical assistance: a comprehensive analysis. ANZ J Surg. Published online August 21, 2023.

20. Grewal H, Dhillon G, Monga V, et al. Radiology Gets Chatty: The ChatGPT Saga Unfolds. Cureus. 2023;15(6):e40135. Published online June 8, 2023.

21. Caruccio L, Cirillo S, Polese G, et al. Can ChatGPT provide intelligent diagnoses? A comparative study between predictive models and ChatGPT to define a new medical diagnostic bot. Expert Syst. Appl.. 2024;235:121186.

22. Rahsepar AA, Tavakoli N, Kim GHJ, et al. How AI Responds to Common Lung Cancer Questions: ChatGPT vs Google Bard. Radiology. 2023;307:e230922.

23. Sarangi PK, Narayan RK, Mohakud S, et al. Assessing the Capability of ChatGPT, Google Bard, and Microsoft Bing in Solving Radiology Case Vignettes. Indian J Radiol Imaging. 2023. Published online December 29, 2023.

