## Supplementary figures and images for "A Comparative Study: Diagnostic Performance of ChatGPT 3.5, Google Bard, Microsoft Bing, and Radiologists in Thoracic Radiology Cases"

### 2-Supplemental Digital Content 2.doc

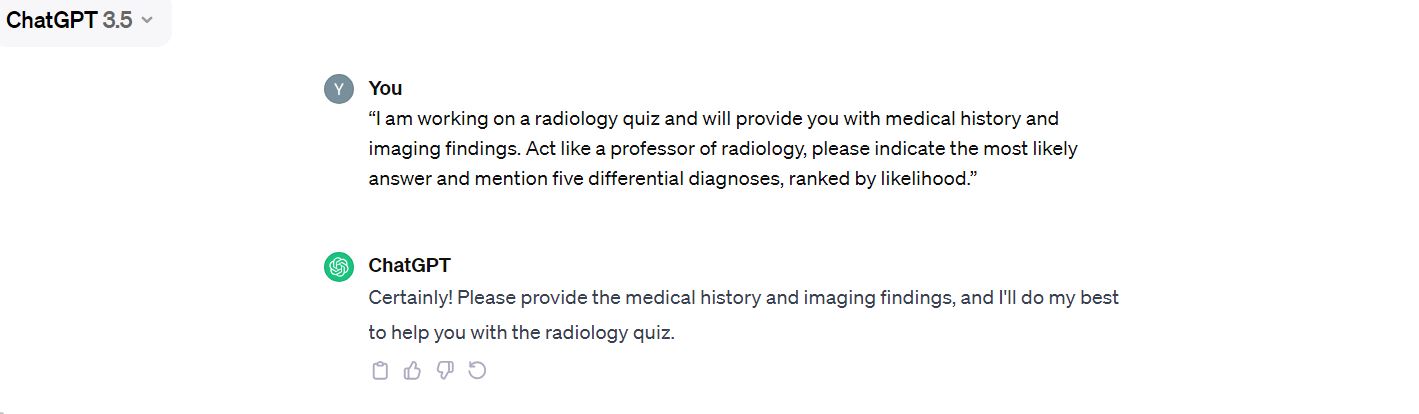
 **CHATGPT**

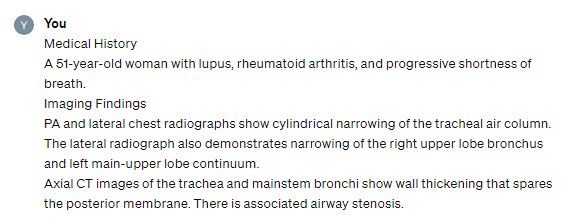

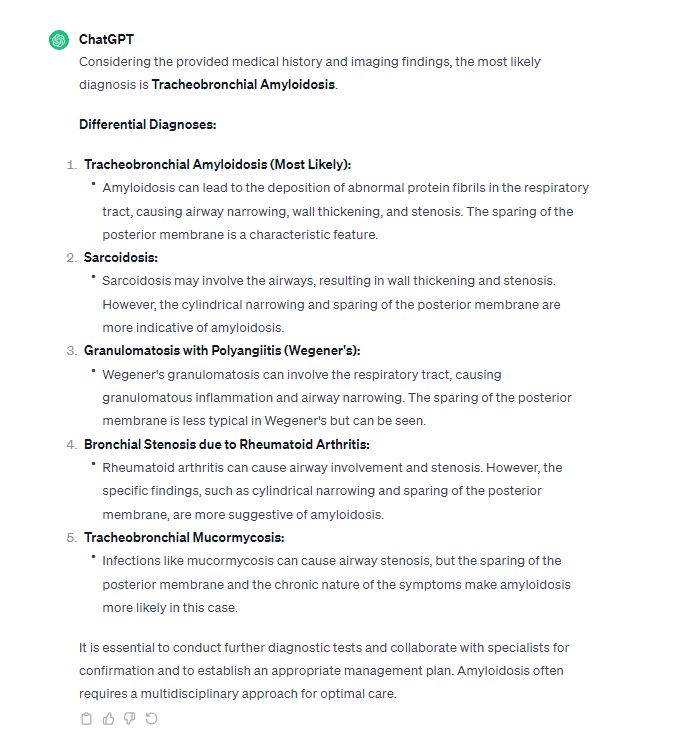

**Microsoft Bing**

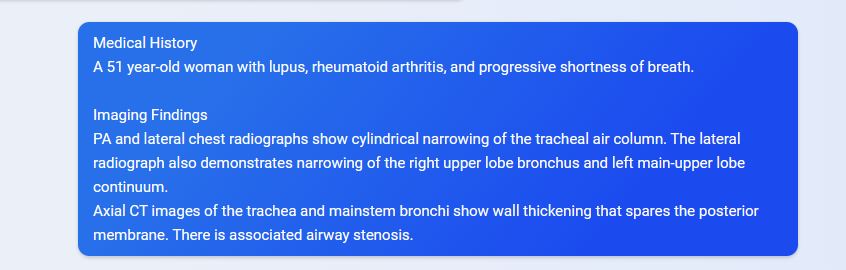

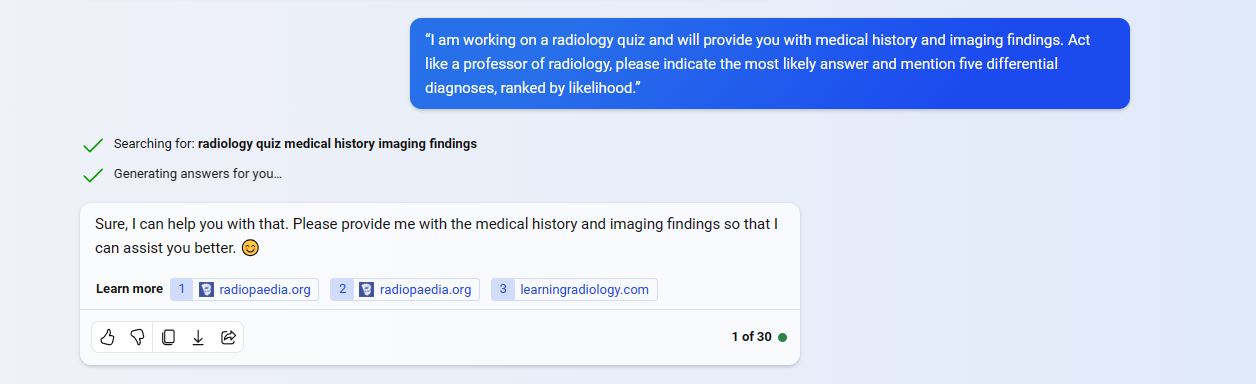

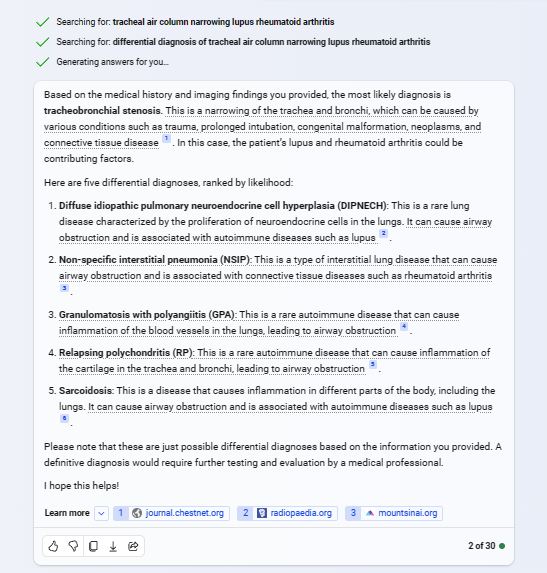

**Google Bard**

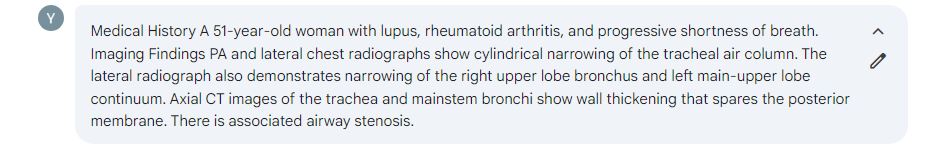

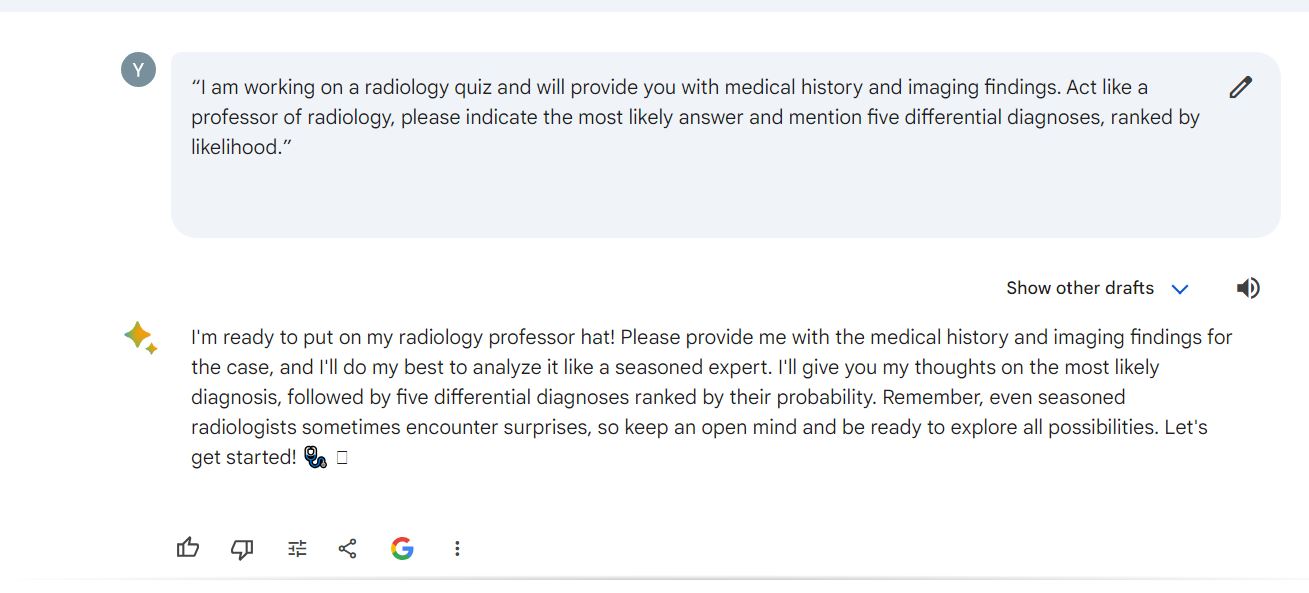

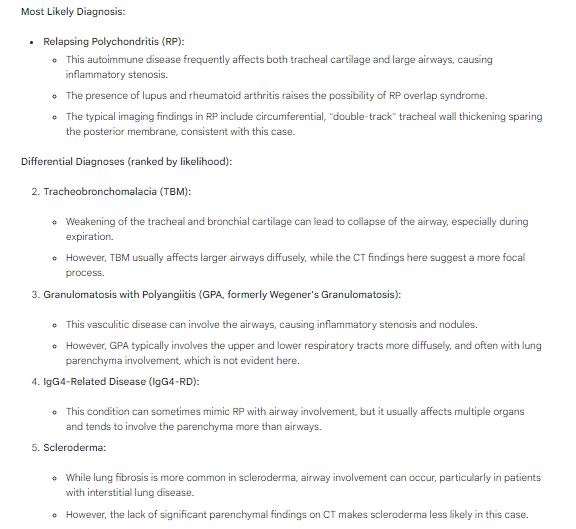
